# Injured worker outcomes after compensation system overhaul: an interrupted time series study

**DOI:** 10.1101/2023.01.13.23284453

**Authors:** Tyler J Lane, Michael Di Donato, Alex Collie

## Abstract

**Objective:** In 2015, South Australia replaced its workers’ compensation system with the aim of improving return to work rates. We tested whether time off work among injured workers changed under the new system, as well as indicators of potential mechanisms.

**Methods:** We conducted a controlled interrupted time series using workers’ compensation claims data. The primary outcome was mean weeks of compensated disability duration. Secondary outcomes tested explanatory mechanisms: 1) claim volumes to determine whether the new system changed the makeup of claimants, and 2) mean employer report and insurer decision times to evaluate whether there had been changes in claim processing. Outcomes were aggregated into monthly units. South Australia was compared to six other Australian workers’ compensation systems. To test for moderation by condition type, disease claims were compared to injury claims and mental health claims to physical health claims.

**Results:** Disability duration and insurer decision time steadily declined before the RTW Act came into effect, but flatlined afterwards. Claim volumes did not change significantly. Employer report time initially increased but gradually decreased until it was lower than the counterfactual. There were non-significant increases in disability durations among injury claims compared to disease claims, and mental health claims compared to physical claims.

**Conclusions:** The increase in disability duration after the RTW Act took effect may be attributable to the disruption of implementing a new compensation system or the elimination of provisional liability entitlements that incentivised early decision making and provided early intervention.

**KEY MESSAGES:** *What is already known on this topic:*

- Workers’ compensation systems are a major determinant of injured worker recovery.
- However, the specific mechanisms of how the compensation system influences outcomes are often opaque, which impedes the design of effective systems.

*What this study adds:*

- This interrupted time series study evaluates the effect of a new workers’ compensation system in South Australia, which was designed specifically to improve return to work rates.
- Time off work increased relative to the counterfactual, which paralleled trends in the time for insurers to decide on liability.

*How this study might affect research, practice, or policy:*

- The findings highlight the importance of understanding how compensation systems influence injured workers outcomes and considering the entire claims process when designing new systems.

## INTRODUCTION

Australian employers are required to insure workers against lost wages and medical expenses resulting from occupational injury. Prior to 2015, South Australia’s workers’ compensation system had among the lowest return to work rates and highest employer premiums in Australia, and was also at risk of becoming financially unviable.^1–3^ The *Return to Work Act 2014* (henceforth, the *RTW Act*) replaced, removed, and modified many aspects of the existing system to address these problems.^4^ The new system came into effect on 1 July 2015 with retrospective effect.^2^ The changes are summarised in Supplementary Table 1.

An independent review suggested that return to work rates improved several years after implementation of the *RTW Act*.^3^ The review argued it was plausible that such improvements would only be observed several years after the reforms since the new system “inherited what was uniformly agreed to be an unsatisfactory scheme” (p. 66). However, it also cautioned that causal attributions with lagged effects is inherently tenuous. Additionally, the analysis was based on summary statistics provided by the new regulator, which were aggregated on an annual basis and lacked granularity better suited for pinpointing effects.

We previously observed that workers’ compensation system reform was followed by an increase in claim processing time,^5,6^ which is predictive of delayed return to work.^7–9^ Longer insurer decision times are also an obstacle to early intervention, which is associated with better outcomes and shorter disability durations.^10–12^ Increases in return to work or shorter disability durations may be due to mechanisms other than improved service delivery. For instance, removing barriers to system access (e.g., lowering or eliminating waiting or employer excess periods) lowers the average injury severity of those who receive compensation. The reduction in disability duration would therefore be due to a change in the compensated cohort rather than an actual improvement in recovery.^13^ As a consequence, policymakers and researchers may come to incorrect conclusions about what mechanisms influence injured worker outcomes and apply changes elsewhere.

Injured worker responses to system change may also vary by the nature of the injured workers’ condition. Trajectories among workers with harder-to-diagnose and less visible conditions like non-specific low back pain tend to be more sensitive to compensation system settings,^14,15^ while different factors affect disability duration among physical and mental health claims.^16^

In this study, we address the following research questions:

1. Did a major compensation system reform reduce disability duration in South Australia?
2. Were there changes in claim volumes, employer report time, or insurer decision time?
3. Were these effects moderated by condition type?

## METHODS

### Setting

There are 11 major workers’ compensations systems in Australia: one for each of the six states and two territories, and three commonwealth schemes: Comcare for federal government and large inter-state private employers; Seacare for seafaring workers; and the Department of Veterans’ affairs system for military employers. This paper only includes the state and territory systems and Comcare, which provide or regulate coverage for 94% of Australia’s workforce.^17^

### Data

We extracted data on claims with ≥1 day of compensated time off work lodged between January 2013 and June 2017 from the *National Data Set for Compensation-based Statistics*.^18^ After identifying issues with self-insurer claims data (see Appendix), data were limited to scheme-insured claims.

### Outcomes

The main study outcome is mean disability duration, operationalised as cumulative compensated weeks off work. While this underestimates the true time to return to work, it is nevertheless considered the most accurate indicator when using administrative claims data.^19^ Individual records were capped at one year to standardise follow-up across claims and to exclude the effect of the two-year cap introduced with the *RTW Act*. Such forced benefit cessation is a poor indicator of actual return to work.

Secondary outcomes focus on alternative mechanisms that could influence disability duration either by changing who gets into the compensation system or the experience of the claims process. Claim volumes were used to test for changes to who gets into the compensation system, while mean days employers took to report an injury/lodge a claim and insurers to accept a claim were used to test for changes to the claims process. To account for skew from extreme outliers, individual records were capped at 60 days for both outcomes. Records with negative durations were assumed to be in error and recoded to zero.

### Condition comparison

To test for moderating effects of condition, claims were grouped into the following for subgroup analyses: 1) disease versus injury claims and 2) mental health versus physical health claims. These are derived from Major Group categories in the Type of Occurrence Classification System 3.1 (TOOCS 3.1).^20^ Major Groups *A* through *G* are classed as *injuries* and *H* through *R* as *diseases and conditions* (shortened to *diseases* to avoid confusion). Diseases include musculoskeletal and connective tissue diseases; digestive system diseases; skin and subcutaneous tissues diseases; nervous system and sense organ diseases; respiratory system diseases; circulatory system diseases; infectious and parasitic diseases; neoplasms (cancer); and other diseases. Injuries include intracranial injuries; fractures; wounds, lacerations, amputations, and internal organ damage; burns; injury to nerves and spinal cord; traumatic joint/ligament and muscle/tendon injury; and other injuries. Mental health conditions (TOOCS Major Group I) are excluded from this analysis and instead separately compared to all physical claims, both injury and disease.

### Analysis

We used an interrupted time series study design with generalised least squares (GLS) regression models for mean duration outcomes (insurer decision, employer report, and disability duration) and negative binomial models for claim volumes. For estimates of the *RTW Act’s* effects on South Australia as a whole, we used a location-based control, the rest of Australia, to account for potential sources of history bias. To determine whether any jurisdictions in the rest of Australia violated the parallel trends assumption or had obvious structural breaks that would make them unsuitable as a comparator,^21^ we plotted outcome trends in the pre-*Act* study period (Supplementary figure 1); none stood out. New South Wales was excluded from the control because there were no data on date employers were notified of injury, which is necessary to calculate employer report time. The *rest of Australia* comparator was therefore comprised of Victoria, Queensland, Western Australia, Tasmania, Australian Capital Territory, and Comcare. To estimate whether condition type modified the *RTW Act’s* effects, comparisons were conducted entirely within South Australia. Disease claim effects were measured relative to injury claims and mental health claims relative to physical claims. To make data comparable, claim volumes in the control group were indexed to the exposure groups using pre-*RTW Act data*.

To account for seasonality, we tested six pairs of sine and cosine terms for both the exposure and control group (24 seasonality terms in total), which were retained only if significant at p ≤ .05.^22^ GLS model outcomes were log-transformed to derive percentage rather than absolute changes and keep all values positive.^23,24^ In GLS models, we tested for residual autocorrelation with autocorrelation and partial autocorrelation plots up to lag 12, which were adjusted using ARMA (Auto-Regressive Moving Average) terms.^25^ All plausible ARMA models were compared using the Akaike Information Criterion to select the one with best fit relative to the number of terms.^22^ Effects from both GLS and negative binomial models are reported as percent changes to the intercept (acute) and slope of trend lines (gradual).^26^ We used a conservative alpha of .99 to account for multiple comparisons. Results were plotted to inform judgements about how meaningful they are. Sensitivity analyses applied a phase-in that excluded the first six months post-*RTW Act* to account for disruptions attributable to adapting to the new system.

Analyses were conducted in R^27^ using RStudio^28^ with the following packages: *broom*.*mixed*,^29^ *forecast*,^30^ *ggpubr*,^31^ *janitor*,^32^ *lubridate*,^33^ *naniar*,^34^ *nlme*,^35^ *readxl*,^36^ *tidyverse*,^37^ and *zoo*.^38^ Aggregated data and analytical code are archived on a public repository.^39^

### Pre-registration and deviations

While this study was pre-registered on the Open Science Framework,^40^ the actual analysis deviates from the pre-registered approach in several ways. Self-insurer claims were only excluded after analysis revealed data quality issues with these employers (see Appendix). This means we could not use covered worker estimates as a denominator, since they are not broken down by scheme and self-insured employer, and therefore instead of claims per 1,000 covered workers, we analyse claim volumes. Negative binomial models were introduced for the claim volume count outcome, as was more appropriate for this data type. Research questions were edited for clarity. The pre-registered analysis plan referred to discretionary versus non-discretionary conditions, which on reflection was deemed too subjective. We previously specified that we would only conduct disability duration analysis by condition type if there was evidence of cohort or process-shaping effects, but decided to conduct them regardless. We did not pre-specify the conservative alpha. The pre-registered plan was to analyse time loss and medical-only claims, which was later limited to time loss claims only to make the cohorts the same across all analyses. Sensitivity analyses excluding the first six months of the *RTW Act* were added after completion of initial analyses.

## RESULTS

There were *N* = 20,610 eligible claims from South Australia and *N* = 315,546 from the rest of Australia excluding New South Wales. For comparisons by condition group within South Australia only, there were *N* = 5,326 disease claims and *N* = 14,014 injury claims, and *N* = 1,259 mental health claims and *N* = 19,340 physical claims; *N* = 11 claims had missing injury data. The small number of mental health claims, spread across 48 time points (four years * 12 months) means these data are underpowered to detect many effects of the *RTW Act* on this group, though we have presented the results of these analyses regardless.

Table 1 summarises claim rates, employer and insurer decision times, and disability durations across groups before and after the *RTW Act* came into effect.

**Table 1.** Monthly means (standard deviation) for each outcome pre- and post-*Return to Work Act 2014*, by group.

|  | Employer report time (days) |  | Insurer decision time (days) |  | Monthly claim volumes |  | Compensated time off work (weeks) |  |
| --- | --- | --- | --- | --- | --- | --- | --- | --- |
|  | Pre | Post | Pre | Post | Pre | Post | Pre | Post |
| **South Australia** | 11.2 (1.2) | 8.2 (3.3) | 29.8 (3.0) | 29.2 (1.8) | 456.8 (53.8) | 402.0 (39.6) | 6.2 (1.2) | 5.7 (0.4) |
| **Rest of Australia** | 8.5 (0.4) | 7.6 (0.4) | 11.7 (0.5) | 10.9 (0.8) | 6,648.8 (576.8) | 6,498.9 (676.0) | 11.5 (0.3) | 12.0 (0.3) |
| **Disease claims** | 15.4 (3.2) | 11.1 (4.7) | 35.9 (3.0) | 35.9 (2.4) | 121.1 (26.7) | 100.8 (16.8) | 7.6 (1.2) | 7.0 (0.9) |
| **Injury claims** | 9.4 (0.9) | 6.8 (2.8) | 25.8 (2.9) | 24.7 (2.1) | 310.2 (28.6) | 273.7 (29.3) | 4.9 (1.0) | 4.4 (0.4) |
| **Mental health claims** | 15.0 (4.5) | 13.8 (8.5) | 52.1 (4.5) | 50.4 (3.8) | 25.2 (7.6) | 27.2 (5.3) | 15.3 (4.8) | 13.5 (2.3) |
| **Physical health claims** | 11.0 (1.3) | 7.9 (3.2) | 28.5 (2.9) | 27.7 (1.8) | 431.3 (48.0) | 374.5 (37.3) | 5.7 (1.0) | 5.1 (0.4) |

**Table 2.** Effects of the *Return to Work Act 2014*; significant effects at *p* < .01 in bold.

|  | Acute effects (99% CI) | Gradual effects (99% CI) |
| --- | --- | --- |
| *Employer report time* |  |  |
| South Australia (relative to control) | **25.5% (7.1% to 43.9%)** | **-6.8% (-8.1% to -5.5%)** |
| Control (rest of Australia) | -2.4% (-15.4% to 10.5%) | 0.0% (-1.0% to 0.9%) |
| Disease claims (relative to control) | -20.1% (-54.5% to 14.4%) | -0.2% (-2.7% to 2.3%) |
| Control (injury claims) | 20.3% (-3.9% to 44.4%) | **-6.9% (-8.6% to -5.1%)** |
| Mental health claims (relative to control) | 70.3% (-9.9% to 150.5%) | -1.9% (-7.7% to 3.9%) |
| Control (physical claims) | 17.4% (-39.4% to 74.1%) | **-6.8% (-10.9% to -2.7%)** |
| *Insurer decision time* |  |  |
| South Australia (relative to control) | **18.5% (7.9% to 29.1%)** | **1.7% (1.0% to 2.5%)** |
| Control (rest of Australia) | -1.5% (-9.0% to 6.0%) | -0.9% (-1.5% to -0.4%) |
| Disease claims (relative to control) | **-17.7% (-32.4% to -2.9%)** | -0.2% (-1.2% to 0.9%) |
| Control (injury claims) | **20.5% (10.1% to 30.9%)** | 0.6% (-0.2% to 1.3%) |
| Mental health claims (relative to control) | **-14.9% (-28.6% to -1.3%)** | 0.6% (-0.4% to 1.6%) |
| Control (physical claims) | **16.7% (7.1% to 26.4%)** | 0.6% (-0.1% to 1.3%) |
| *Claim volumes* |  |  |
| South Australia (relative to control) | 7.8% (-4.0% to 19.6%) | 0.6% (-0.2% to 1.4%) |
| Control (rest of Australia) | 0.5% (-8.1% to 9.1%) | 0.5% (-0.1% to 1.0%) |
| Disease claims (relative to control) | **25.9% (3.4% to 48.4%)** | 1.2% (-0.4% to 2.8%) |
| Control (injury claims) | -1.2% (-16.9% to 14.4%) | 0.6% (-0.6% to 1.7%) |
| Mental health claims (relative to control) | 43.5% (-0.8% to 88.0%) | 2.6% (-0.5% to 5.7%) |
| Control (physical claims) | 5.1% (-26.5% to 36.6%) | 0.9% (-1.4% to 3.1%) |
| *Disability duration* |  |  |
| All claims (relative to control) | **26.5% (14.6% to 38.4%)** | **2.4% (1.6% to 3.3%)** |
| Control (rest of Australia) | -0.3% (-8.4% to 7.7%) | -0.1% (-0.7% to 0.5%) |
| Disease claims (relative to control) | -18.6% (-43.6% to 6.3%) | -0.3% (-2.0% to 1.5%) |
| Control (injury claims) | **22.7% (5.5% to 39.8%)** | **1.8% (0.6% to 3.0%)** |
| Mental health claims (relative to control) | 15.0% (-16.6% to 46.6%) | **2.4% (0.2% to 4.7%)** |
| Control (physical claims) | 16.2% (-6.0% to 38.4%) | **1.9% (0.3% to 3.5%)** |

### Employer report

Employer report time in South Australia increased 25.5% (99% CI: 7.1% to 43.9%) after the *RTW Act* came into effect, followed by a 6.8% per month gradual reduction (99% CI: -8.1% to -5.5%) that brought it well below the counterfactual (see Figure 1).

**Figure 1.**
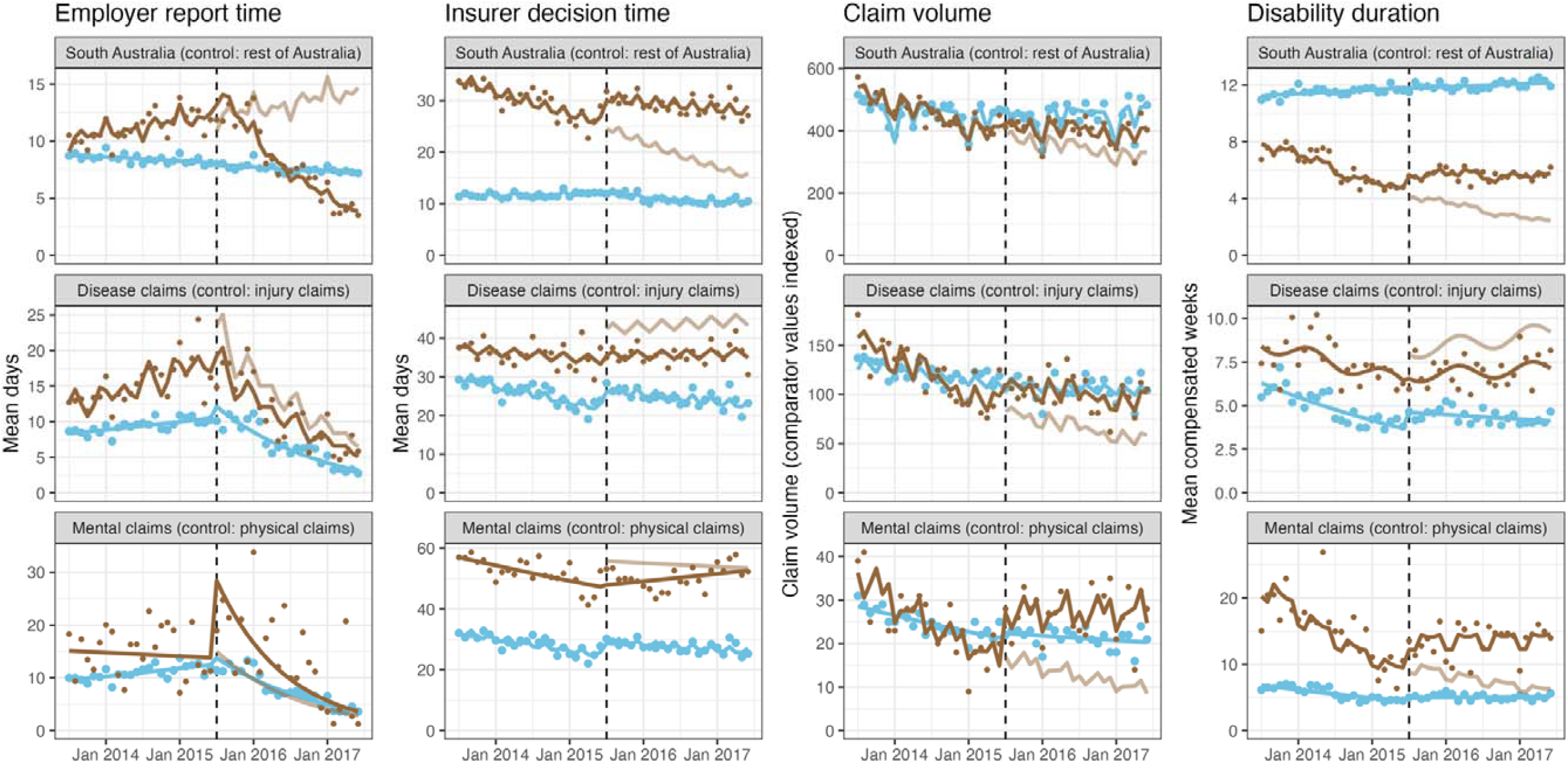
Time series plots of the *RTW Act*’s effects; brown = exposure, blue = control; faded brown = counterfactual (expected trajectory if *RTW Act* were never implemented)

There were significant gradual reductions in the injury (−6.9%; 99% CI: -8.6% to -5.1%) and physical claims (−6.8%; 99% CI: -10.9% to -2.7%) control groups. Disease and mental health claims did not significantly differ their controls, though plotted results indicate a sharp and substantial increase in employer report time for mental health claims (70.3%; 99% CI: -9.9% to 150.5%), which sharply abated to near the counterfactual.

### Insurer decision time

Insurer decision time increased by 18.5% (99% CI: 7.9% to 29.1%) after the RTW Act was implemented and continued to increase relative to the counterfactual by 1.7% per month (99% CI: 1.0% to 2.5%). The plotted results in Figure 1 suggest this gradual increase is better characterised as stabilising into a steadier reduction after faster declines in previous years.

Both the injury and physical claim control groups saw significant immediate increases in insurer decision time. The plotted results suggest no real immediate effect in disease and mental health claims, though relative their controls, these were significant decreases.

### Claim volumes

Claim volumes increased by 7.8% in South Australia relative to the rest of Australia, though the change was non-significant (−4.0% to 19.6%). Relative to injury claims, disease claims increased 25.9% (99% CI: 3.4% to 48.4%). The plotted results in Figure 1 suggest a real effect. Mental health claims increased 43.5% (−0.8% to 88.0%), and though this was not significant at our conservative alpha of .01, we highlight this finding because it is clearly visible in Figure 1. There were no detectable gradual effects.

### Disability duration

Disability duration in South Australia increased 26.5% (99% CI: 14.6% to 38.4%) after implementation of the *RTW Act*. There was also a 2.4% per month gradual increase (99% CI: 1.6% to 3.3%) relative to the counterfactual trend, though the plotted results suggest disability duration had stabilised after steadily decreasing in the preceding years.

The injury control group increased 22.7% (5.5% to 39.8%) in addition to a 1.8% per month gradual increase (0.6% to 3.0%), which the plotted results in Figure 1 suggest is a stabilisation after a steady decrease. Relative to these, there were no detectable effects in disease claims. But, rather than indicating that disease claims kept pace with injury claims, the plotted results indicate no change in disease claims after the *RTW Act* came into effect. There were no immediate effects in disability duration among mental health or physical claims, though there was a 2.4% per month gradual increase (99% CI: 0.2% to 4.7%) among mental health claims, which is on top of the 1.9% gradual increase in physical claims (99% CI: 0.3% to 3.5%). Figure 1 suggests the gradual effect in mental health claim disability durations is a stabilisation following a decrease in the secular trend.

### Sensitivity analyses

Excluding the first six months post-*RTW Act* changed some effects from significant to non-significant and vice-versa, though there were no meaningful changes to magnitudes or directions of effects. The results of sensitivity analyses are presented in Supplementary figure 4 and Supplementary table 2.

## DISCUSSION

The stated aims of the *RTW Act* were to improve return to work rates among injured workers. We found instead that disability durations increased slightly. Additionally, whereas disability durations had previously been declining, after implementation of the *RTW Act* they stabilised. There is no corresponding change to the control group consisting of the rest of Australia, which suggests that the change observed in South Australia is attributable to the *RTW Act*.

There are several reasons to attribute the increase in disability duration to the increase insurer decision time. First, their post-*RTW Act* patterns match, with stabilisation of what had been a decline up until the *RTW Act*’s implementation. By itself, this would be weak evidence that the changes in insurer decision time and disability duration were causally linked. However, we designated insurer decision time as an outcome of interest a priori^41^ because of its link to disability duration.^7^

This raises the question of how the *RTW Act* could have increased disability duration. One possibility is the administrative burden of adapting to an entirely new compensation system, which we observed following earlier compensation system reforms in Tasmania and South Australia.^6^ The *RTW Act* also eliminated of provisional liability. Under the previous compensation system, workers’ compensation applicants were entitled to wage replacement and medical benefits if the insurer had not decided on liability within seven calendar days. The *RTW Act* increased the waiting period to 10 business days and made payments recoverable if the claim was rejected.^1,2^ This removed an incentive for insurers to make a quick decision on claim liability since they were no longer liable for provisional (and unrecoverable) benefits to applications that they would ultimately reject. Recognising this problem, the Parliamentary Committee tasked with investigating the *RTW Act* recommended reintroducing provisional liability, but only for early intervention services.^1^

Like insurer decision times, employer report times are independently associated with disability duration.^7^ However, the steady reduction in employer report times after implementation of the *RTW Act* did not correspond to a reduction in disability duration. It remains possible that the improvement in employer report time had some effect on disability duration, but it was masked by stronger competing forces. The next question is why employer report time decreased. First, there was an initial spike in employer report time, which may reflect the administrative burden of employers trying to learn a new system. That employer report time fell after this initial spike may indicate that in the long run, the new system was more efficient. For instance, the *RTW Act* marked the first time that employers could lodge claims over the telephone, where previously it was a paper application that had to be posted.^2,3^

Our findings contrast with other investigations of the *RTW Act*’s effects. National Return to Work Survey data found the return to work rate among injured workers 7-9 months post injury was 82% in both 2012/13 and 2013/14 and 81% in 2015/16,^42^ a negligible difference. As noted in the introduction, an independent review found return to work rates only increased several years after the *RTW Act* passed.^3^

The reasons for contrasting results need to be addressed. One issue is the difference in how return to work is measured. We converted claims data into monthly time series and used a continuous measure of time off work (mean compensated time off work), while the Return to Work Survey and independent review used dichotomous measures (back at work at points in time). Our use of time series better accounted for secular trends, such as the steady reduction in disability duration before the *RTW Act* came into effect, while the continuous disability duration measure is more sensitive to individual variations in time off work than dichotomous return to work at points int time. Further, our monthly aggregations were more granular and therefore better able to detect effects^26^ than the biennial data in the Return to Work Survey and annual data in the Independent Review.

It remains possible that in the long run, the *RTW Act* reduced time off work for injured workers. The reduction in employer report time is a particularly noteworthy long-term improvement, which, after some time, might have helped injured workers get early intervention and return to work more quickly. However, the trends identified within these analyses appeared fairly stable in the two-year post-Act period that remained even when excluding the first six months to account for any disruption. Later improvements would likely deviate considerably from the trends established with our analyses and would be difficult to attribute to the *RTW Act*.

### Differences by condition

Contrary to expectations, we found no evidence that the *RTW Act* affected disability duration among disease claims. Instead, we found a substantial increase in the control group, injury claims. Similarly, there was an increase in insurer decision times among injuries but not diseases. This suggests that the rise in disability durations generally is largely attributable to an increase in the injury subgroup. In the above section, offered two explanations for the increase in insurer decision time: administrative burden of adopting to a new compensation system and elimination of provisional liability. What remains unclear is why such a mechanism would only affect injury claims, which tend to result from acute trauma that is more easily diagnosed and attributed to work, and less vulnerable the diagnostic and attribution problems in disease conditions.

Among mental health claims, there was a non-significant increase in employer report time – which quickly abated, claim volumes, and disability duration. There was no immediate effect on insurer decision time, but there was a gradual increase. As above, these effects are difficult to explain. While non-significant, the substantial increase in claim volumes was most surprising since the *RTW Act* introduced a theoretically stricter criterion for mental health claims: for mental health claims employment must be “*the significant* contributing cause of the injury” but for physical claims only “*a* significant contributing cause” (emphasis added). As described elsewhere, we would expect an increase in claim volumes to reflect a system that has become easier to access, reducing both the average injury severity (since milder injuries are now more likely to become a claim) and the iatrogenic effects of a strict system.^8,13^

### Strengths and limitations

Study strengths include the use of a powerful quasi-experimental study design, the interrupted time series, as well as population-level data, controls to account for history bias and to directly compare effects by condition, and testing for specific mechanisms that may explain changes in disability duration. In addition, the study was pre-registered with each deviation noted in this paper, and we archived aggregated study data and cleaning and analytical code on a public repository.

Limitations include the difficulty of distinguishing causal mechanisms due competing factors within a comprehensive system change. Cumulative compensated time loss data served as a proxy for disability duration/return to work, which systematically underestimates true disability duration.^43^ Small numbers in mental health claims mean that while some effects appeared obvious from plotted results, statistical tests remained non-significant. Due to the unique nature of Australian compensation systems, the findings may not generalise to other contexts.

## CONCLUSIONS

South Australia introduced the *RTW Act* to overhaul its workers’ compensation system and improve injured worker outcomes. Instead, we found that while disability durations were decreasing before the *RTW Act* took effect, it flatlined afterwards. There is reason to attribute this to a simultaneous flatlining of insurer decision time, which in turn was likely caused either by the burden of adapting to a new compensation system or the elimination of provisional liability. Faster insurer decisions and earlier intervention may be simple ways to improve injured worker outcomes.

## Data Availability

All data produced are available online at https://doi.org/10.26180/21625778. We only provide aggregated data that were analysed in the study, as individual-level were potentially identifiable. We do provide cleaning code to demonstrate how data were aggregated.

## APPENDIX PROBLEMS WITH CLAIMS DATA FROM SELF-INSURED EMPLOYERS

After the *RTW Act* was implemented, there was a substantial increase in claims with missing injury data from self-insured employers. This is illustrated in Supplementary figure 1. Post-*RTW Act*, self-insured claims with missing injury data had 69.6% longer employer report times and 33.3% shorter insurer decision times (both *p* ≤ .001); there was no detectable difference in disability duration (*p* = 0.530). See Supplementary figure 2 for a distribution of outcomes. As a result, analyses were limited to scheme-insured claims.

**Supplementary Figure 1.**
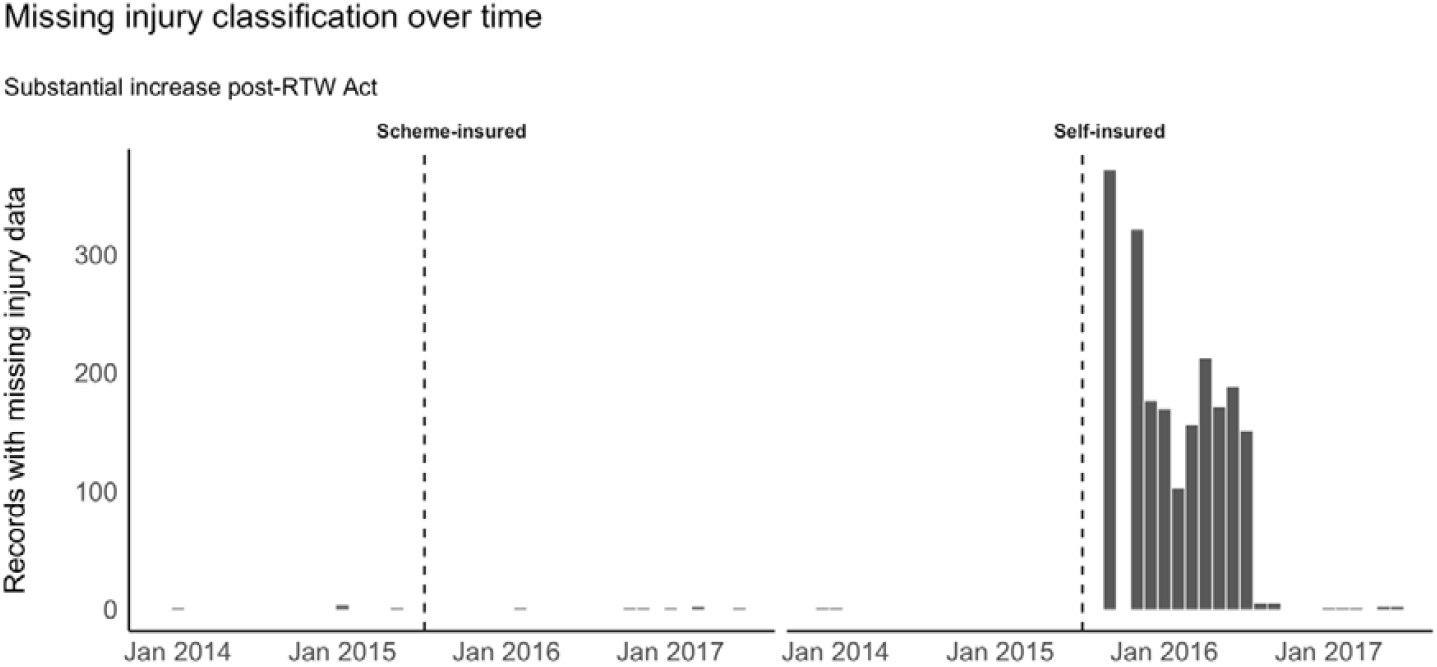
Monthly count of claims with missing data, scheme and self-insured employers.

**Supplementary Figure 2.**
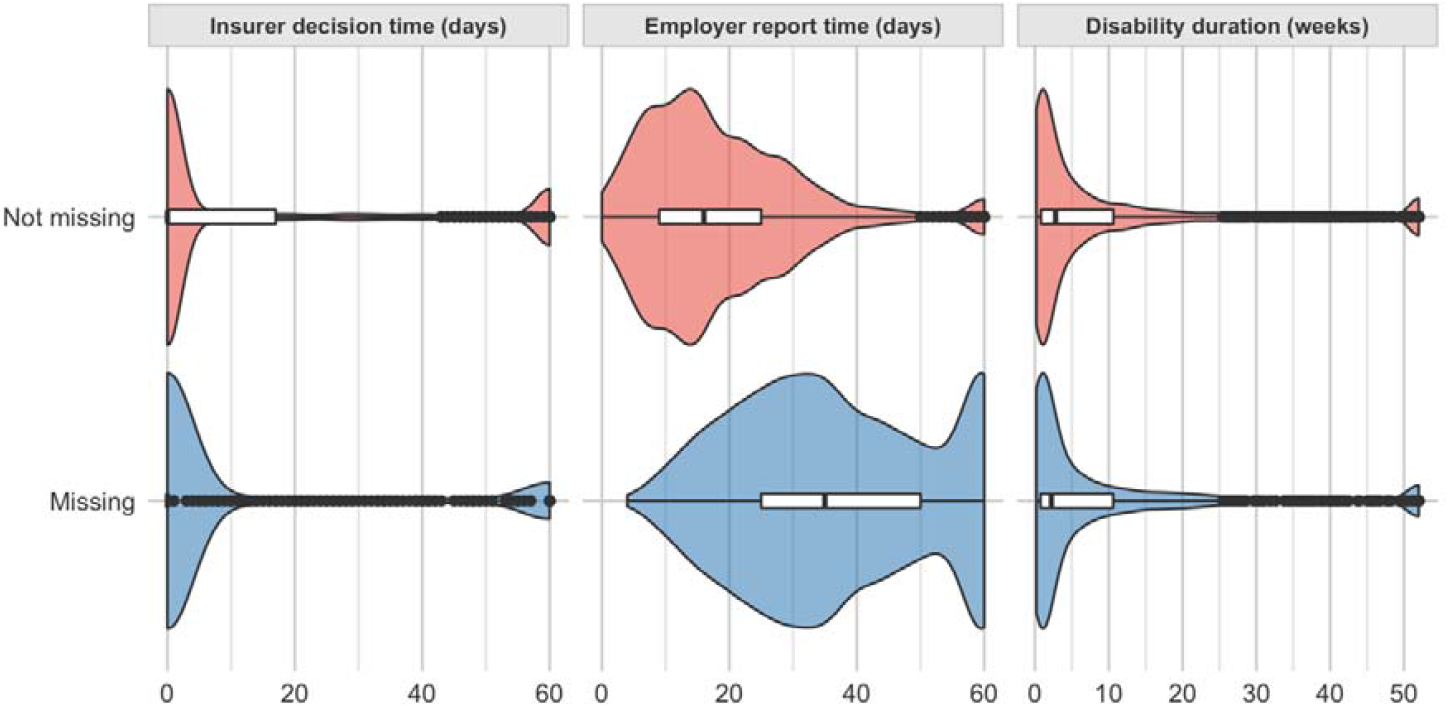
Distribution in outcomes between claims with missing and non-missing injury data from self-insured employers, post-*RTW Act*.

## ADDITIONAL FIGURES

**Supplementary Figure 3.**
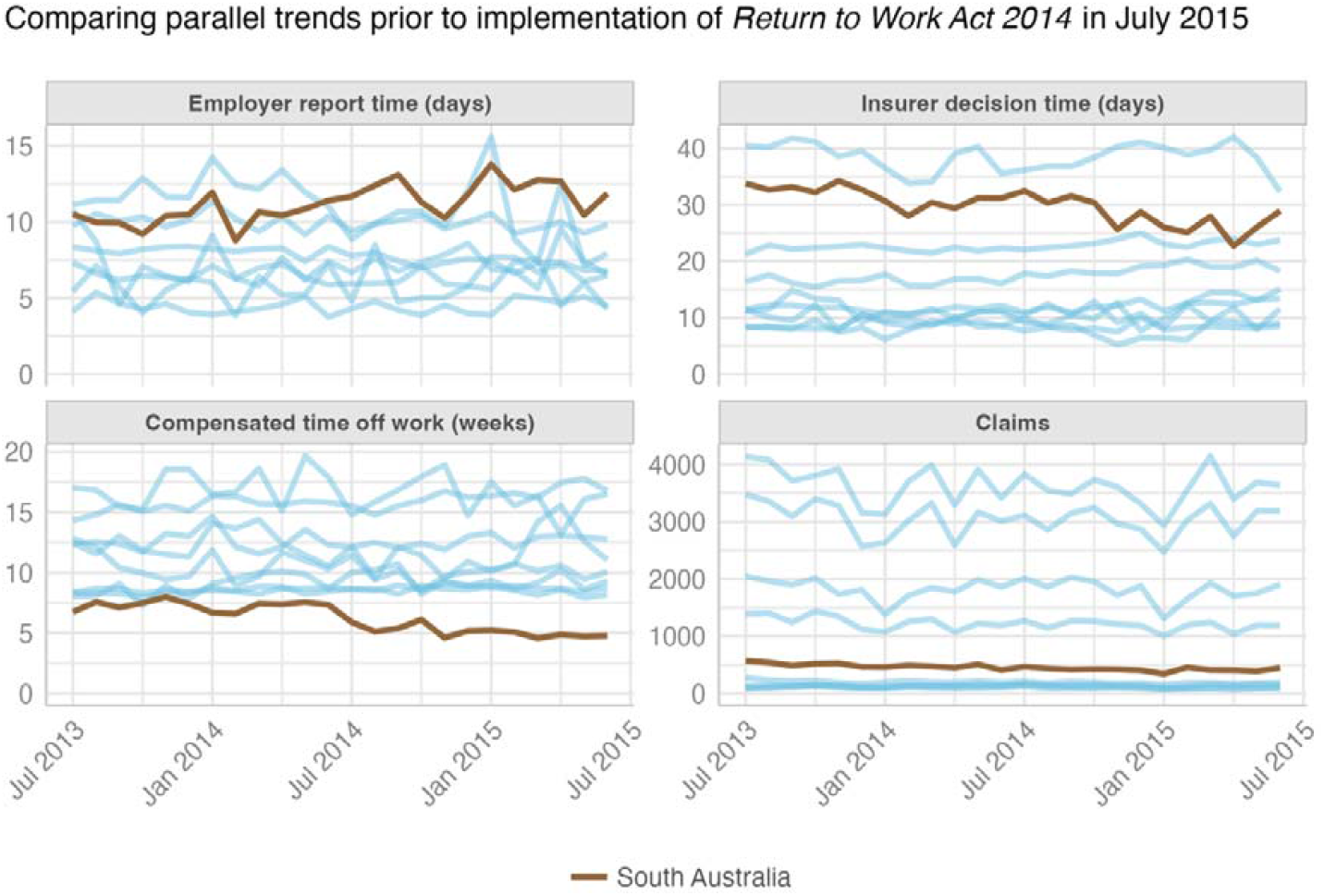
**Pre-RTW Act trends in outcomes between South Australia and other jurisdictions (blue); the plots show no evidence that the parallel trends assumption, a key component of interrupted time series analysis, has been violated, except disability duration in Victoria (top blue line in bottom chart)**

**Supplementary Figure 4.**
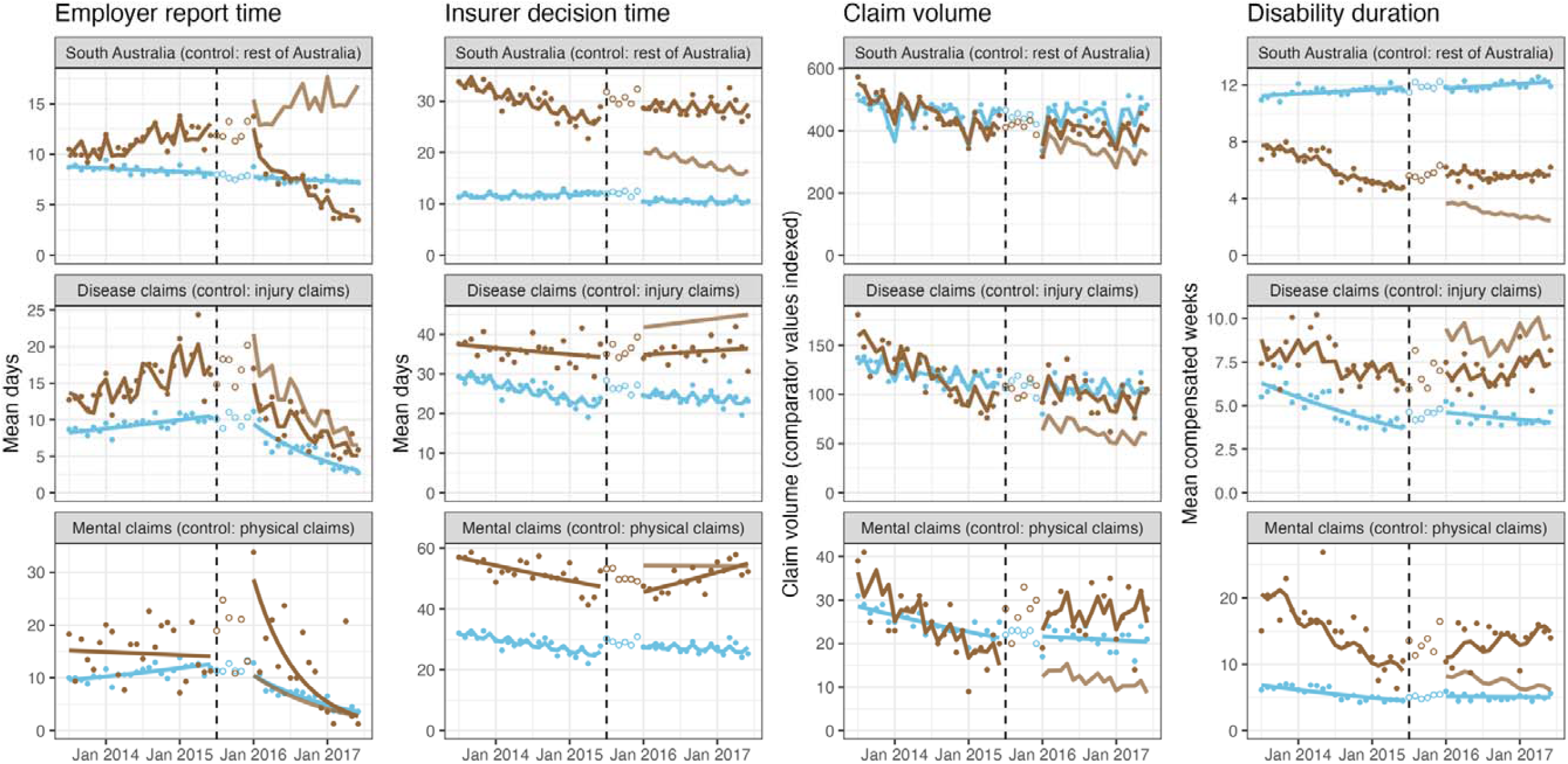
**Time series plots of the *RTW Act*’s effects from sensitivity analyses that excluded the first six months after implementation; brown = exposure, blue = control; faded brown = counterfactual (expected trajectory if *RTW Act* were never implemented); hollow points were excluded from analyses**

## CONTRIBUTIONS

AC conceived the study. TJL conducted analyses and drafted the manuscript, with input from MDD and AC. All authors approved the final manuscript.

## ACKNOWLEDGEMENTS

The authors would like to thank representatives of Return to Work South Australia and Safe Work Australia for their comments on an earlier draft of this manuscript.

## ETHICS

This study received ethics approval from the Monash University Human Research Ethics Committee (CF14/2995 – 2014001663)

---

**Supplementary Table 1.**
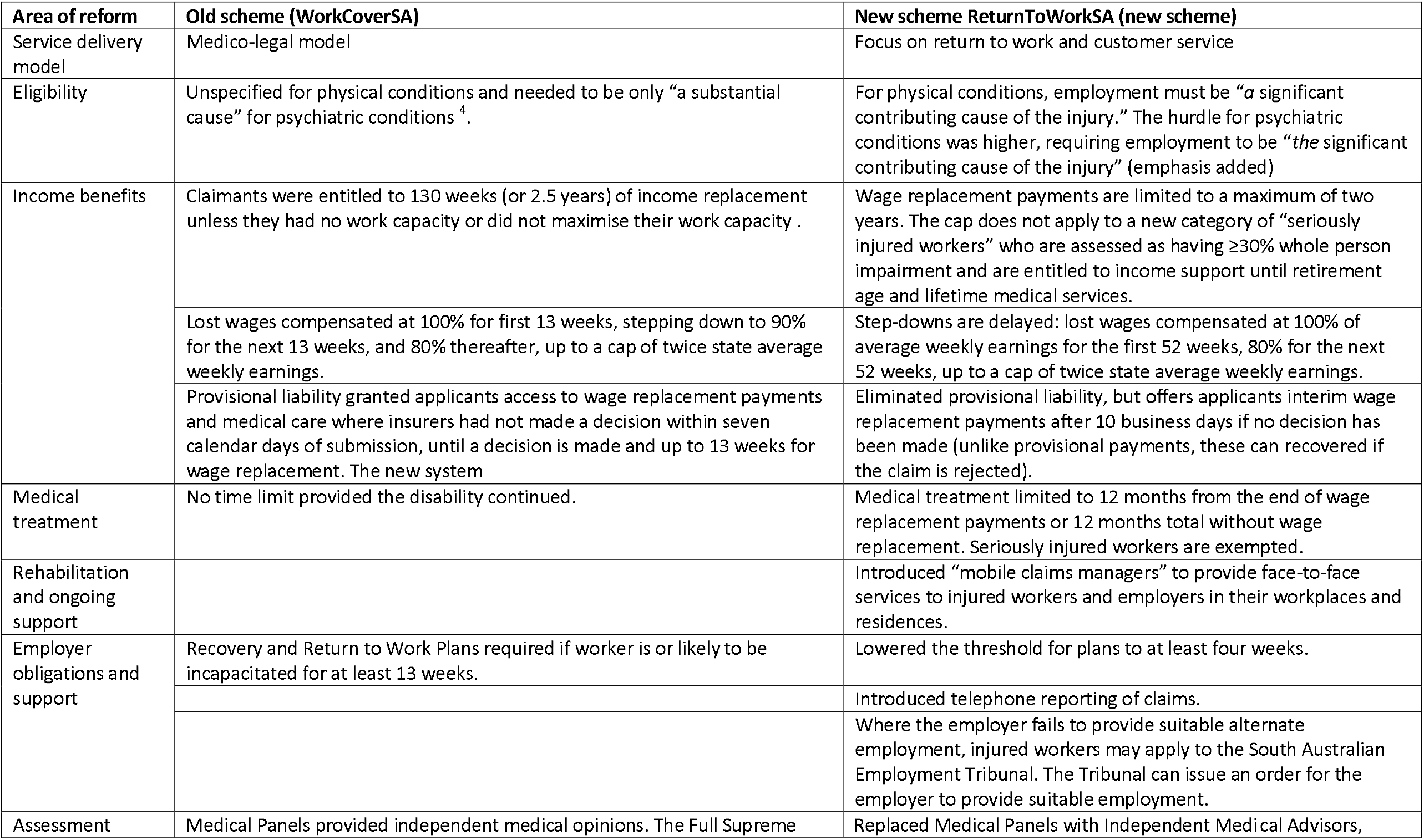
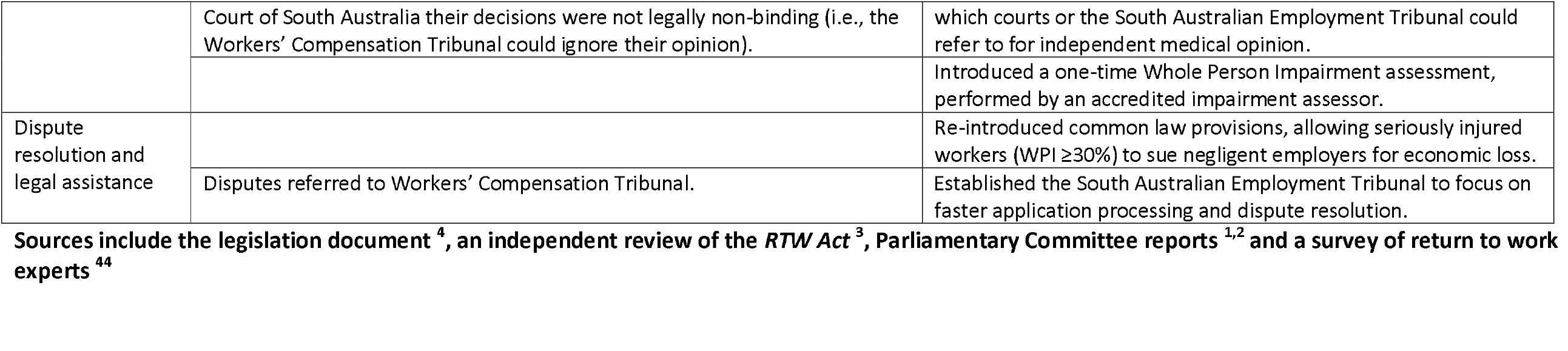
Major changes to the South Australian workers’ compensation system with the implementation of the *Return to Work Act 2014*.

**Supplementary table 2.** Effects of the *Return to Work Act 2014* from sensitivity analyses that excluded first six months after implementation; significant effects at *p* < .01 in bold.

|  | Acute effects (99% CI) | Gradual effects (99% CI) |
| --- | --- | --- |
| *Employer report time* |  |  |
| South Australia (relative to control) | **32.9% (9.2% to 56.7%)** | **-7.6% (-9.2% to -6.1%)** |
| Control (rest of Australia) | -1.5% (-18.2% to 15.1%) | -0.1% (-1.2% to 1.0%) |
| Disease claims (relative to control) | **-48.2% (-95.5% to -0.8%)** | 1.2% (-1.8% to 4.2%) |
| Control (injury claims) | 37.5% (4.4% to 70.7%) | **-7.9% (-10.0% to -5.7%)** |
| Mental health claims (relative to control) | **136.7% (21.6% to 251.8%)** | -5.6% (-13.0% to 1.8%) |
| Control (physical claims) | 26.5% (-54.9% to 107.9%) | **-7.3% (-12.6% to -2.1%)** |
| *Insurer decision time* |  |  |
| South Australia (relative to control) | **25.1% (10.5% to 39.6%)** | **1.4% (0.4% to 2.3%)** |
| Control (rest of Australia) | **-12.3% (-22.6% to -2.0%)** | -0.3% (-1.0% to 0.3%) |
| Disease claims (relative to control) | -17.1% (-39.8% to 5.7%) | -0.2% (-1.6% to 1.3%) |
| Control (injury claims) | **16.8% (0.6% to 32.9%)** | 0.8% (-0.2% to 1.8%) |
| Mental health claims (relative to control) | **-24.9% (-44.2% to -5.5%)** | 1.1% (-0.1% to 2.4%) |
| Control (physical claims) | 13.5% (-0.2% to 27.2%) | 0.8% (-0.1% to 1.7%) |
| *Claim volumes* |  |  |
| South Australia (relative to control) | 5.5% (-11.6% to 22.6%) | 0.7% (-0.4% to 1.8%) |
| Control (rest of Australia) | 0.9% (-11.1% to 13.0%) | 0.4% (-0.4% to 1.1%) |
| Disease claims (relative to control) | **35.0% (1.8% to 68.3%)** | 0.7% (-1.4% to 2.9%) |
| Control (injury claims) | -5.8% (-29.2% to 17.5%) | 0.8% (-0.7% to 2.3%) |
| Mental health claims (relative to control) | 44.7% (-19.4% to 109.2%) | 2.5% (-1.5% to 6.6%) |
| Control (physical claims) | 4.0% (-43.2% to 50.2%) | 0.9% (-2.1% to 3.9%) |
| *Disability duration* |  |  |
| All claims (relative to control) | **26.0% (7.6% to 44.4%)** | **2.4% (1.2% to 3.6%)** |
| Control (rest of Australia) | -2.2% (-15.1% to 10.7%) | 0.0% (-0.8% to 0.9%) |
| Disease claims (relative to control) | **-34.8% (-69.4% to -0.1%)** | 0.6% (-1.6% to 2.9%) |
| Control (injury claims) | **27.3% (2.8% to 51.7%)** | **1.5% (0.0% to 3.1%)** |
| Mental health claims (relative to control) | 3.7% (-45.3% to 52.8%) | 3.0% (-0.2% to 6.1%) |
| Control (physical claims) | 17.5% (-16.9% to 51.8%) | 1.8% (-0.4% to 4.0%) |

## The RECORD statement – checklist of items, extended from the STROBE statement, that should be reported in observational studies using routinely collected health data

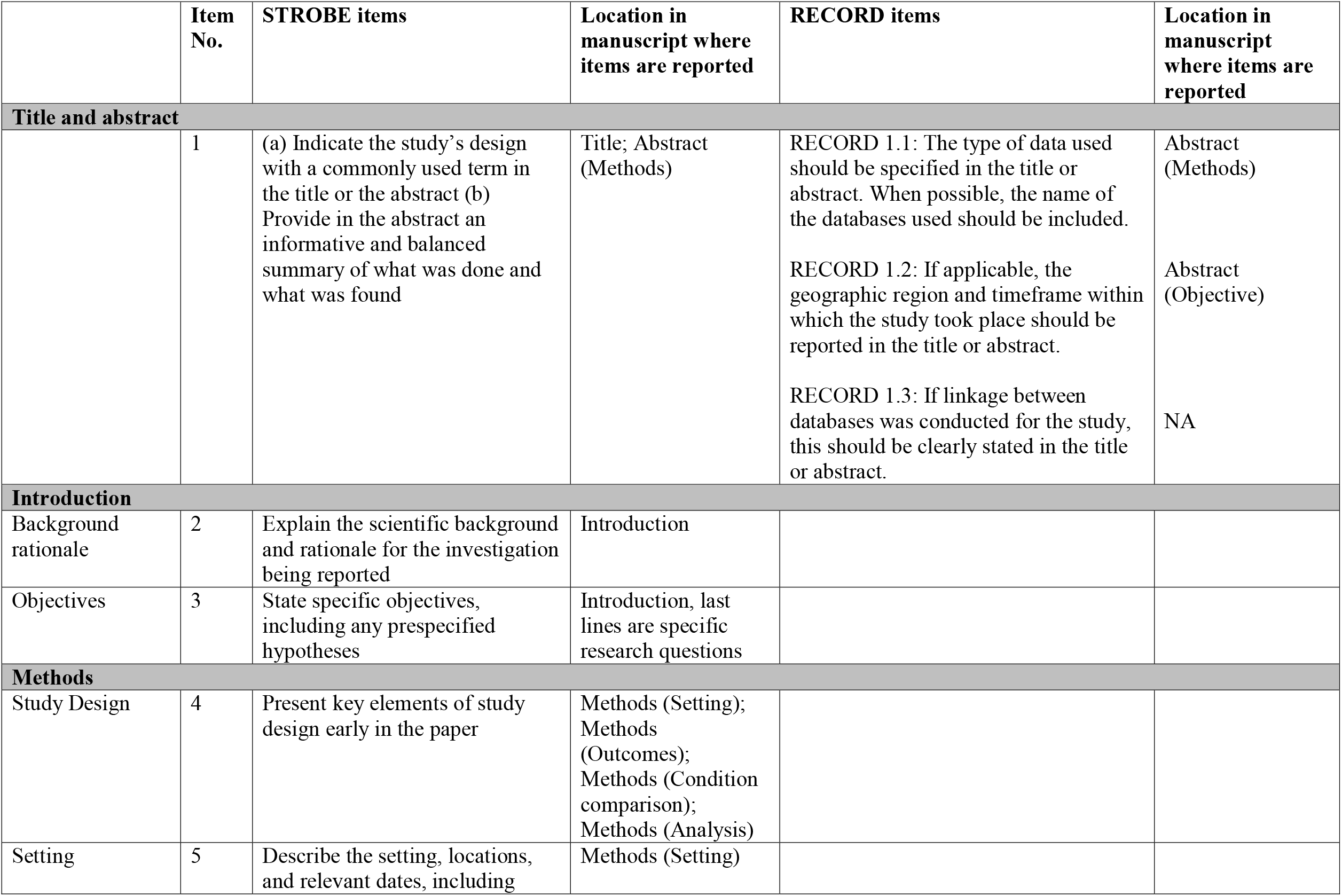
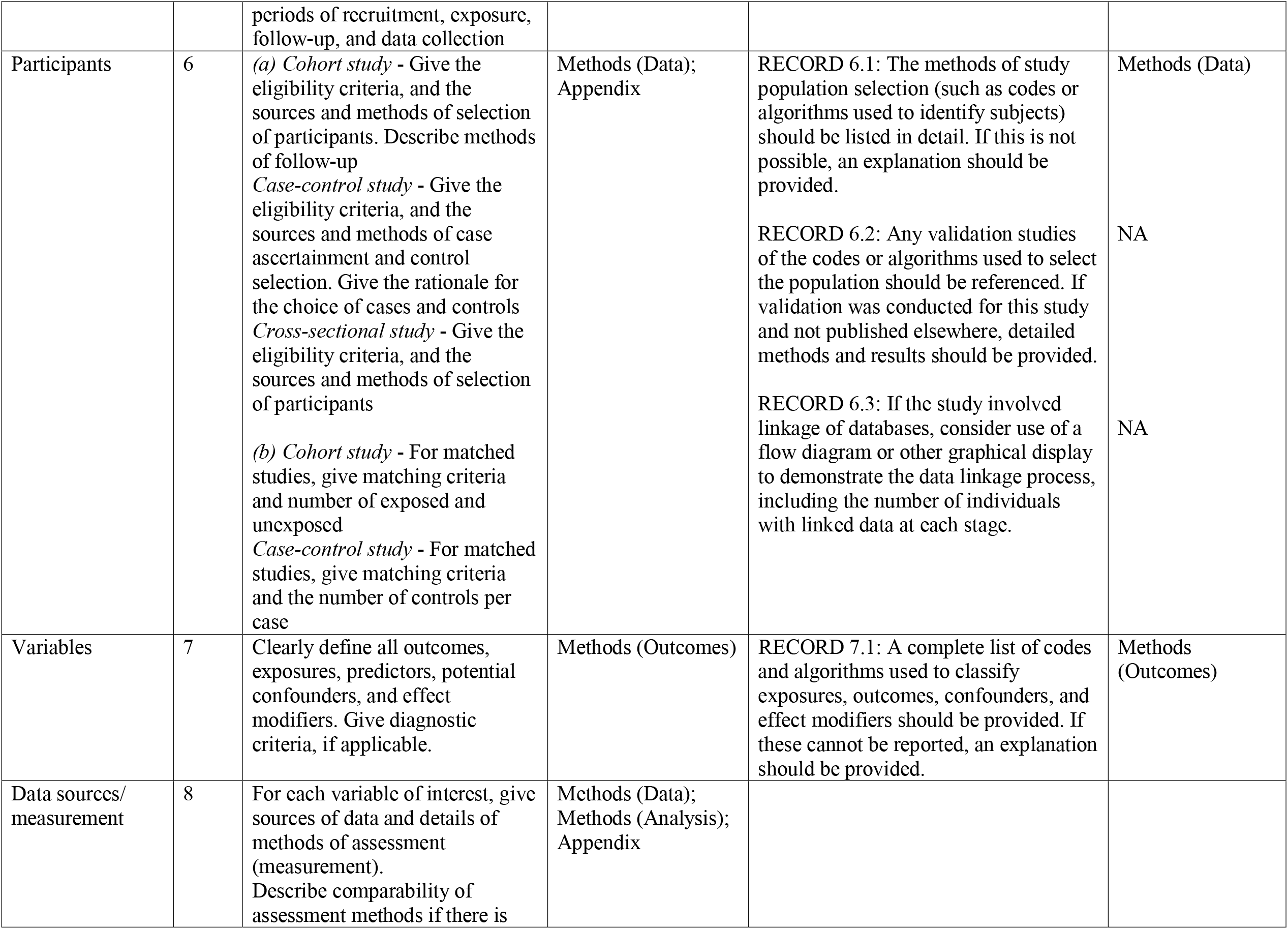
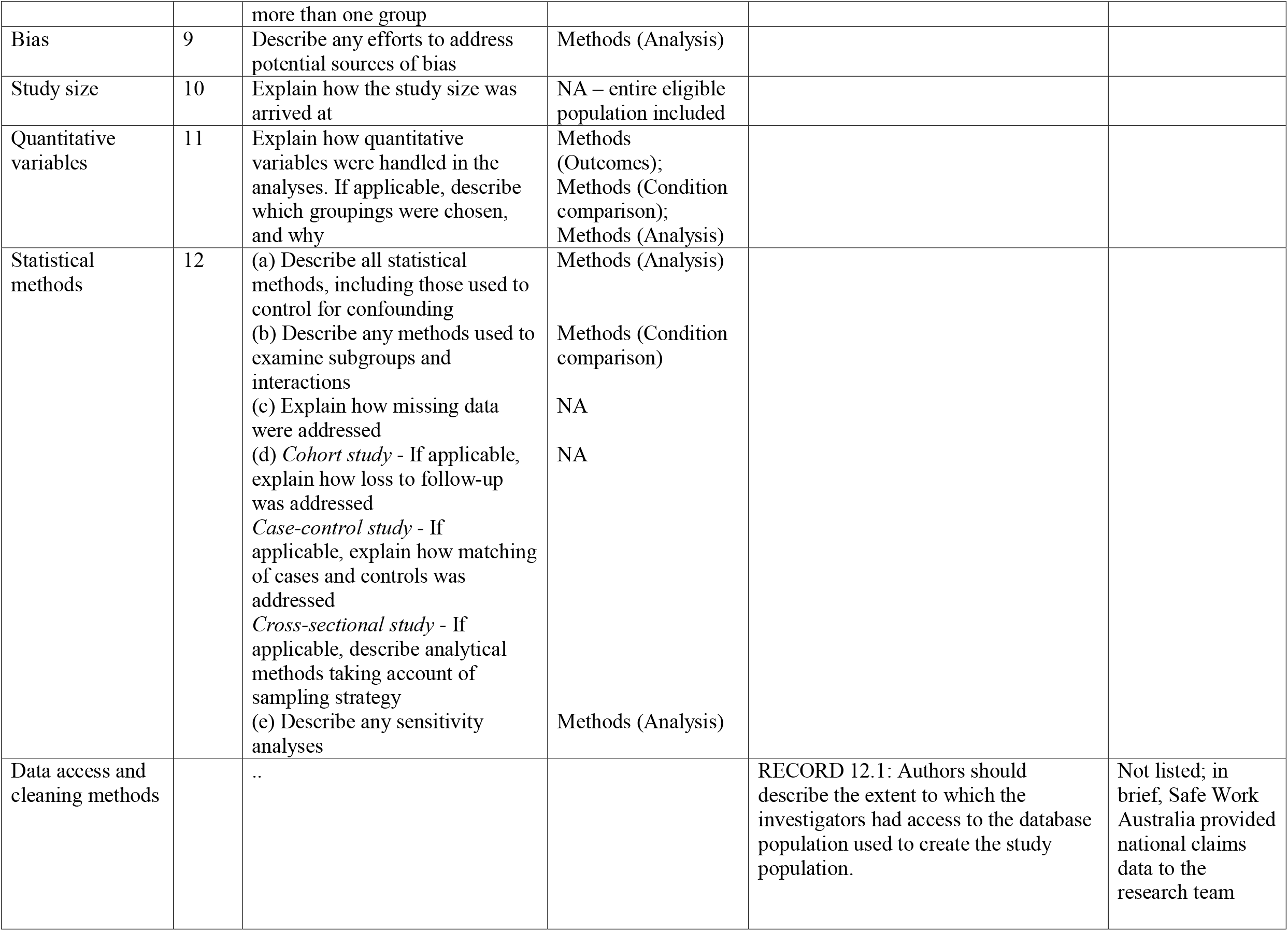
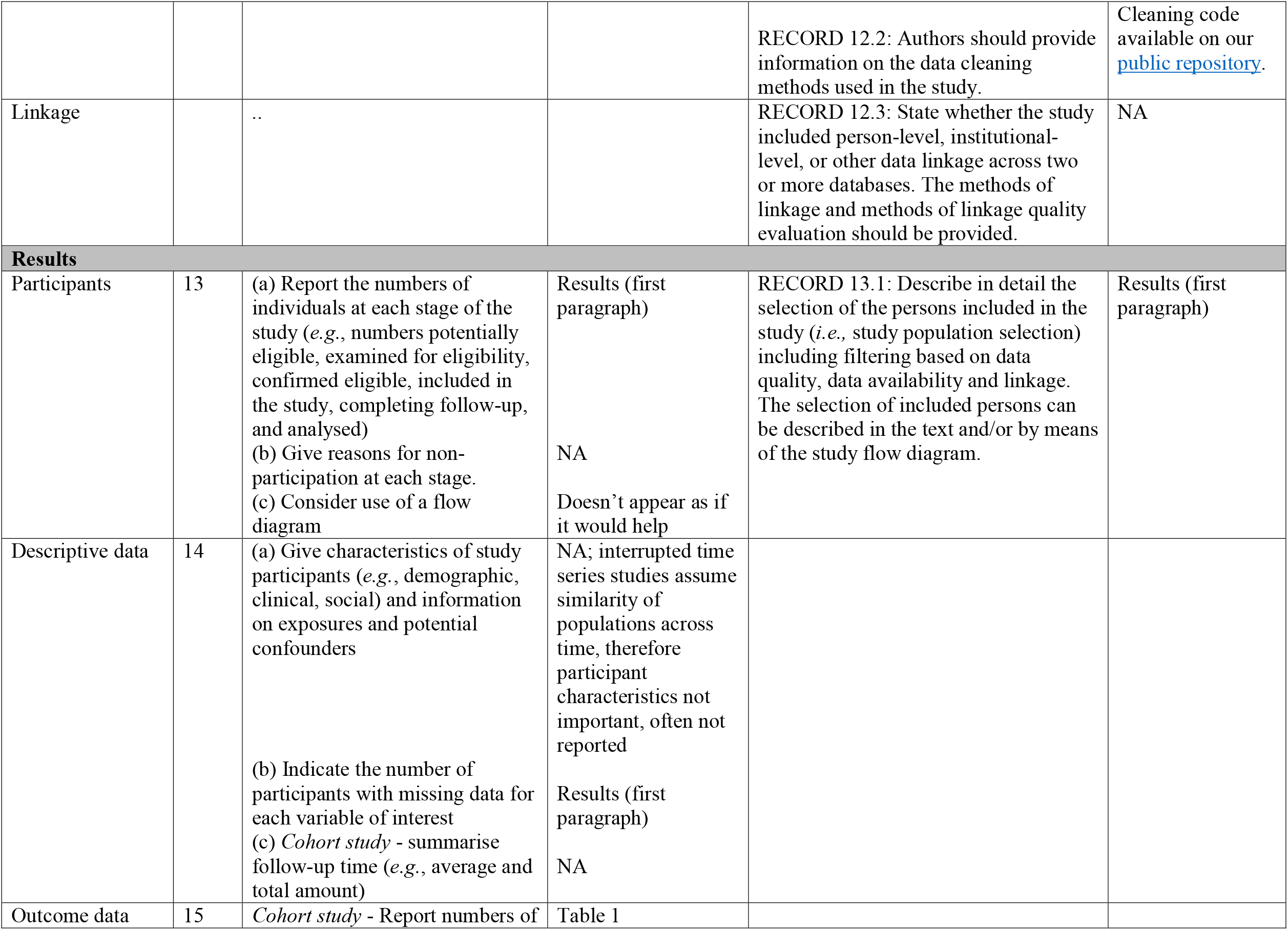
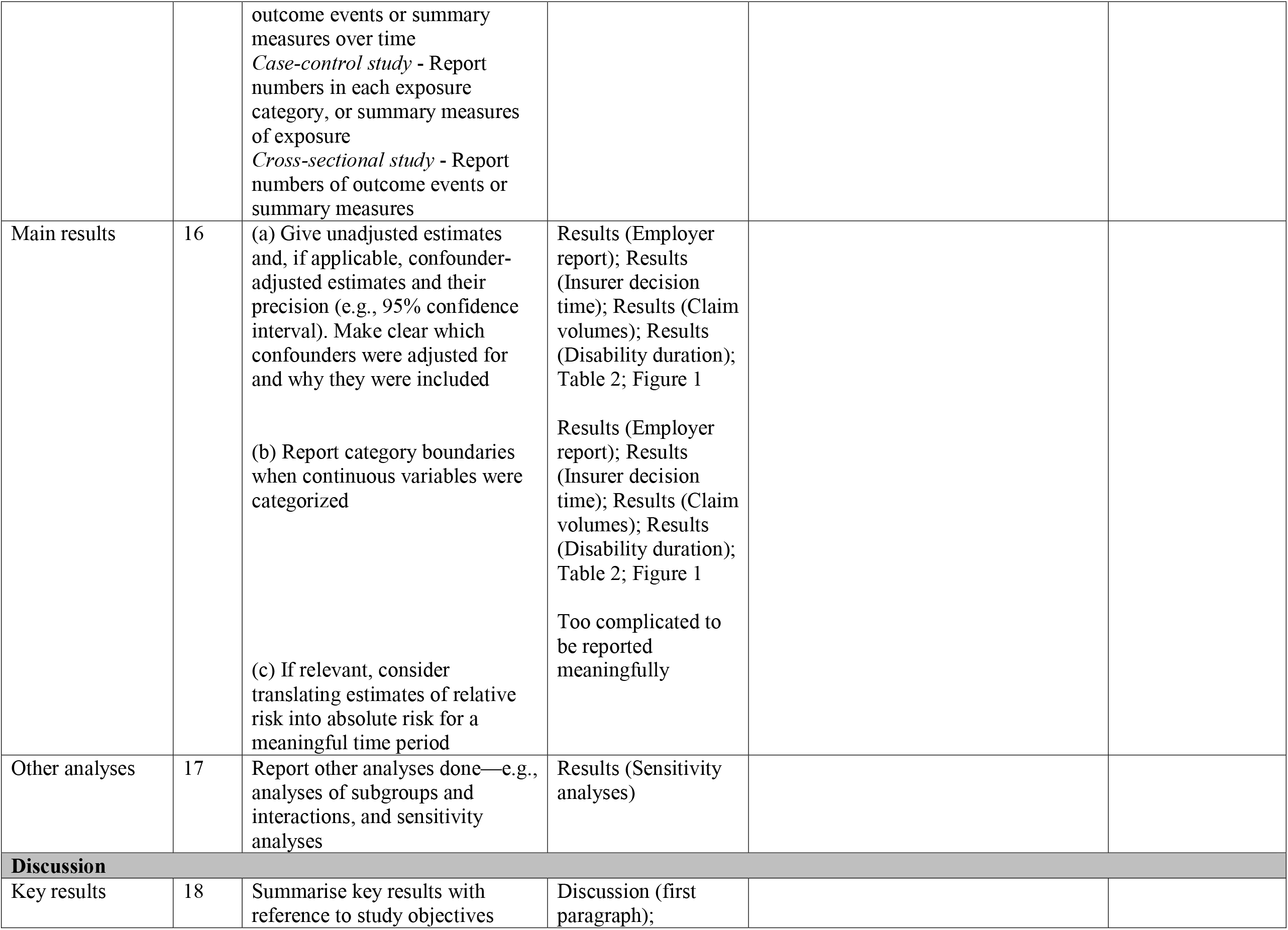
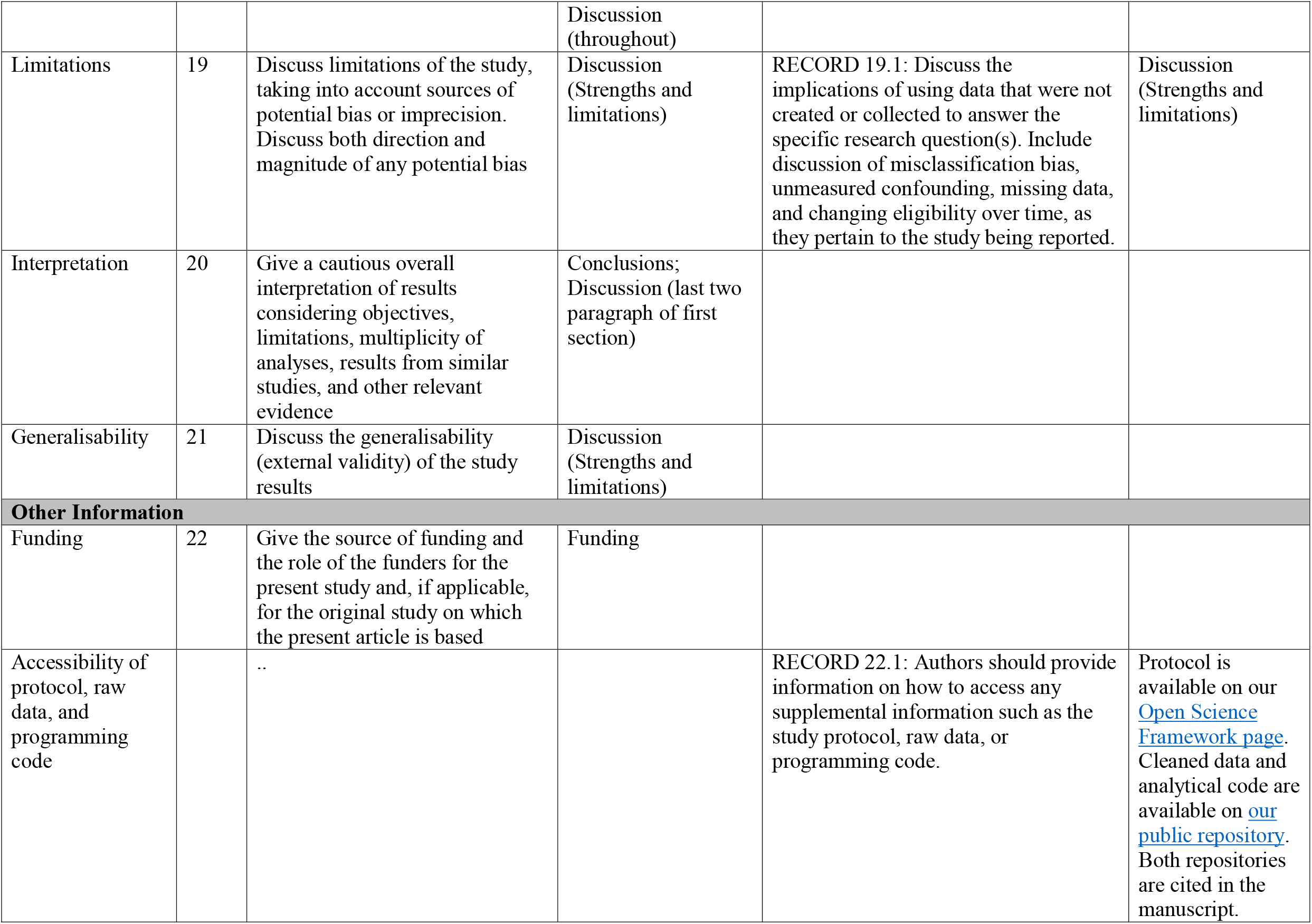
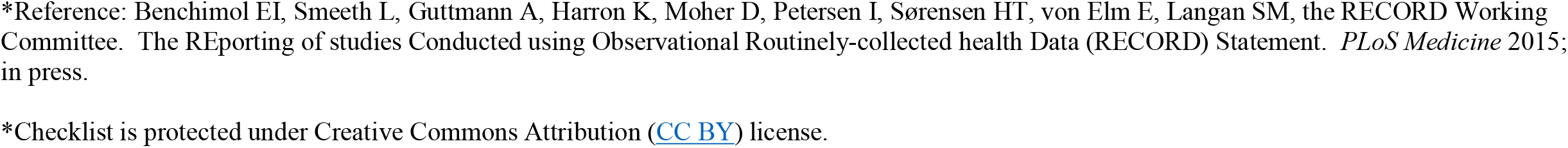

|  | Item No. | STROBE items | Location in manuscript where items are reported | RECORD items | Location in manuscript where items are reported |
| --- | --- | --- | --- | --- | --- |
| **Title and abstract** |  |  |  |  |  |
|  | 1 | (a) Indicate the study's design with a commonly used term in the title or the abstract (b) Provide in the abstract an informative and balanced summary of what was done and what was found | Title; Abstract (Methods) | RECORD 1.1: The type of data used should be specified in the title or abstract. When possible, the name of the databases used should be included.<br>RECORD 1.2: If applicable, the geographic region and timeframe within which the study took place should be reported in the title or abstract.<br>RECORD 1.3: If linkage between databases was conducted for the study, this should be clearly stated in the title or abstract. | Abstract (Methods)<br>Abstract (Objective)<br>NA |
| **Introduction** |  |  |  |  |  |
| Background rationale | 2 | Explain the scientific background and rationale for the investigation being reported | Introduction |  |  |
| Objectives | 3 | State specific objectives, including any prespecified hypotheses | Introduction, last lines are specific research questions |  |  |
| **Methods** |  |  |  |  |  |
| Study Design | 4 | Present key elements of study design early in the paper | Methods (Setting);<br>Methods (Outcomes);<br>Methods (Condition comparison);<br>Methods (Analysis) |  |  |
| Setting | 5 | Describe the setting, locations, and relevant dates, including periods of recruitment, exposure, follow-up, and data collection | Methods (Setting) |  |  |
| Participants | 6 | (a) *Cohort study* - Give the eligibility criteria, and the sources and methods of selection of participants. Describe methods of follow-up<br>*Case-control study* - Give the eligibility criteria, and the sources and methods of case ascertainment and control selection. Give the rationale for the choice of cases and controls<br>*Cross-sectional study* - Give the eligibility criteria, and the sources and methods of selection of participants<br>(b) *Cohort study* - For matched studies, give matching criteria and number of exposed and unexposed<br>*Case-control study* - For matched studies, give matching criteria and the number of controls per case | Methods (Data); Appendix | RECORD 6.1: The methods of study population selection (such as codes or algorithms used to identify subjects) should be listed in detail. If this is not possible, an explanation should be provided.<br>RECORD 6.2: Any validation studies of the codes or algorithms used to select the population should be referenced. If validation was conducted for this study and not published elsewhere, detailed methods and results should be provided.<br>RECORD 6.3: If the study involved linkage of databases, consider use of a flow diagram or other graphical display to demonstrate the data linkage process, including the number of individuals with linked data at each stage. | Methods (Data)<br>NA<br>NA |
| Variables | 7 | Clearly define all outcomes, exposures, predictors, potential confounders, and effect modifiers. Give diagnostic criteria, if applicable. | Methods (Outcomes) | RECORD 7.1: A complete list of codes and algorithms used to classify exposures, outcomes, confounders, and effect modifiers should be provided. If these cannot be reported, an explanation should be provided. | Methods (Outcomes) |
| Data sources/<br>measurement | 8 | For each variable of interest, give sources of data and details of methods of assessment (measurement). Describe comparability of assessment methods if there is more than one group | Methods (Data);<br>Methods (Analysis);<br>Appendix |  |  |
| Bias | 9 | Describe any efforts to address potential sources of bias | Methods (Analysis) |  |  |
| Study size | 10 | Explain how the study size was arrived at | NA – entire eligible population included |  |  |
| Quantitative variables | 11 | Explain how quantitative variables were handled in the analyses. If applicable, describe which groupings were chosen, and why | Methods (Outcomes);<br>Methods (Condition comparison);<br>Methods (Analysis) |  |  |
| Statistical methods | 12 | (a) Describe all statistical methods, including those used to control for confounding<br>(b) Describe any methods used to examine subgroups and interactions<br>(c) Explain how missing data were addressed<br>(d) *Cohort study* - If applicable, explain how loss to follow-up was addressed<br>*Case-control study* - If applicable, explain how matching of cases and controls was addressed<br>*Cross-sectional study* - If applicable, describe analytical methods taking account of sampling strategy<br>(e) Describe any sensitivity analyses | Methods (Analysis)<br>Methods (Condition comparison)<br>NA<br>NA<br>Methods (Analysis) |  |  |
| Data access and cleaning methods |  | .. |  | RECORD 12.1: Authors should describe the extent to which the investigators had access to the database population used to create the study population. | Not listed; in brief, Safe Work Australia provided national claims data to the research team |
|  |  |  |  | RECORD 12.2: Authors should provide information on the data cleaning methods used in the study. | Cleaning code available on our public repository. |
| Linkage |  | .. |  | RECORD 12.3: State whether the study included person-level, institutional-level, or other data linkage across two or more databases. The methods of linkage and methods of linkage quality evaluation should be provided. | NA |
| **Results** |  |  |  |  |  |
| Participants | 13 | (a) Report the numbers of individuals at each stage of the study (*e.g.*, numbers potentially eligible, examined for eligibility, confirmed eligible, included in the study, completing follow-up, and analysed)<br>(b) Give reasons for non-participation at each stage.<br>(c) Consider use of a flow diagram | Results (first paragraph)<br>NA<br>Doesn't appear as if it would help | RECORD 13.1: Describe in detail the selection of the persons included in the study (*i.e.*, study population selection) including filtering based on data quality, data availability and linkage. The selection of included persons can be described in the text and/or by means of the study flow diagram. | Results (first paragraph) |
| Descriptive data | 14 | (a) Give characteristics of study participants (*e.g.*, demographic, clinical, social) and information on exposures and potential confounders<br>(b) Indicate the number of participants with missing data for each variable of interest<br>(c) *Cohort study* - summarise follow-up time (*e.g.*, average and total amount) | NA; interrupted time series studies assume similarity of populations across time, therefore participant characteristics not important, often not reported<br>Results (first paragraph)<br>NA |  |  |
| Outcome data | 15 | *Cohort study* - Report numbers of<br>outcome events or summary measures over time<br>*Case-control study* - Report numbers in each exposure category, or summary measures of exposure<br>*Cross-sectional study* - Report numbers of outcome events or summary measures | Table 1 |  |  |
| Main results | 16 | (a) Give unadjusted estimates and, if applicable, confounder-adjusted estimates and their precision (e.g., 95% confidence interval). Make clear which confounders were adjusted for and why they were included<br>(b) Report category boundaries when continuous variables were categorized<br>(c) If relevant, consider translating estimates of relative risk into absolute risk for a meaningful time period | Results (Employer report); Results (Insurer decision time); Results (Claim volumes); Results (Disability duration); Table 2; Figure 1<br>Results (Employer report); Results (Insurer decision time); Results (Claim volumes); Results (Disability duration); Table 2; Figure 1<br>Too complicated to be reported meaningfully |  |  |
| Other analyses | 17 | Report other analyses done—e.g., analyses of subgroups and interactions, and sensitivity analyses | Results (Sensitivity analyses) |  |  |
| **Discussion** |  |  |  |  |  |
| Key results | 18 | Summarise key results with reference to study objectives | Discussion (first paragraph); |  |  |
|  |  |  | Discussion (throughout) |  |  |
| Limitations | 19 | Discuss limitations of the study, taking into account sources of potential bias or imprecision. Discuss both direction and magnitude of any potential bias | Discussion (Strengths and limitations) | RECORD 19.1: Discuss the implications of using data that were not created or collected to answer the specific research question(s). Include discussion of misclassification bias, unmeasured confounding, missing data, and changing eligibility over time, as they pertain to the study being reported. | Discussion (Strengths and limitations) |
| Interpretation | 20 | Give a cautious overall interpretation of results considering objectives, limitations, multiplicity of analyses, results from similar studies, and other relevant evidence | Conclusions; Discussion (last two paragraph of first section) |  |  |
| Generalisability | 21 | Discuss the generalisability (external validity) of the study results | Discussion (Strengths and limitations) |  |  |
| **Other Information** |  |  |  |  |  |
| Funding | 22 | Give the source of funding and the role of the funders for the present study and, if applicable, for the original study on which the present article is based | Funding |  |  |
| Accessibility of protocol, raw data, and programming code |  | .. |  | RECORD 22.1: Authors should provide information on how to access any supplemental information such as the study protocol, raw data, or programming code. | Protocol is available on our Open Science Framework page. Cleaned data and analytical code are available on our public repository. Both repositories are cited in the manuscript. |

\*Reference: Benchimol EI, Smeeth L, Guttman A, Harron K, Moher D, Petersen I, Sørensen HT, von Elm E, Langan SM, the RECORD Working Committee. The REporting of studies Conducted using Observational Routinely-collected health Data (RECORD) Statement. *PLoS Medicine* 2015; in press.
\*Checklist is protected under Creative Commons Attribution (CC BY) license.

## REFERENCES

1. Parliamentary Committee on Occupational Safety, Rehabilitation and Compensation. Final Report into the Referral for an Inquiry into the Return to Work Act and Scheme. (Parliament of South Australia, 2017).

2. Parliamentary Committee on Occupational Safety, Rehabilitation, and Compensation. Interim Report into the Referral for an Inquiry into the Return to Work Act and Scheme. (Parliament of South Australia, 2017).

3. Mansfield, J. Independent Review of the Return to Work Act 2014. (2018).

4. Parliament of South Australia. Return to Work Act 2014. (2015).

5. Collie, A, Beck, D, Gray, S. & Lane, TJ. Impact of legislative reform on benefit access and disability duration in workers’ compensation: an interrupted time series study. Occup. Environ. Med. 77, 32–39 (2020).

6. Lane, TJ, Gray, S, Hassani-Mahmooei, B & Collie, A. Effectiveness of employer financial incentives in reducing time to report worker injury: an interrupted time series study of two Australian workers’ compensation jurisdictions. BMC Public Health 18, 100 (2018).

7. Gray, SE, Lane, TJ, Sheehan, L & Collie, A. Association between workers’ compensation claim processing times and work disability duration: Analysis of population level claims data. Health Policy 123, 982–991 (2019).

8. Collie, A, Sheehan, L, Lane, TJ, Gray, S & Grant, G. Injured worker experiences of insurance claim processes and return to work: a national, cross-sectional study. BMC Public Health 19, 927 (2019).

9. Besen, E, Harrell, M & Pransky, G. Lag times in reporting injuries, receiving medical care, and missing work: Associations with the length of work disability in occupational back injuries. J. Occup. Environ. Med. 58, 53–60 (2016).

10. Kucera, KL, Lipscomb, HJ, Silverstein, B & Cameron, W. Predictors of delayed return to work after back injury: A case-control analysis of union carpenters in Washington State. Am. J. Ind. Med. 52, 821–830 (2009).

11. Lusted, M. Predicting return to work after rehabilitation for low back injury. Aust. J. Physiother. 39, 203–210 (1993).

12. Stover, B, Wickizer, TM, Zimmerman, F, Fulton-Kehoe, D & Franklin, GM. Prognostic factors of long-term disability in a workers’ compensation system. J. Occup. Environ. Med. 49, 31–40 (2007).

13. Lane, TJ. Collider bias in administrative workers’ compensation claims data: A challenge for cross-jurisdictional research. J. Occup. Rehabil. 32, 161–169 (2022).

14. Butler, RJ, Durbin, D. & Helvacian, NM. Increasing claims for soft tissue injuries in workers’ compensation: Cost shifting and moral hazard. J. Risk Uncertain. 13, 73–87 (1996).

15. Butler, RJ, Gardner, H. & Kleinman, NL. Workers’ Compensation: Occupational Injury Insurance’s Influence on the Workplace. in Handbook of Insurance: Second Edition (ed. Dionne, G) 449–469 (Springer, 2013). doi:10.1007/978-1-4614-0155-1

16. Huijs, JJJM, Koppes, LLJ, Taris, T. & Blonk, RWB. Differences in predictors of return to work among long-term sick-listed employees with different self-reported reasons for sick leave. J. Occup. Rehabil. 22, 301–311 (2012).

17. Lane, TJ, Collie, A & Hassani-Mahmooei, B. Work-related injury and illness in Australia, 2004 to 2014. (Institute for Safety Compensation & Recovery Research, 2016).

18. National Occupational Health and Safety Commission. National Data Set for Compensation-based Statistics 3rd Edition (Revision 1). (Commonwealth of Australia, 2004).

19. Krause, N, Dasinger, LK, Deegan, LJ, Brand, R. & Rudolph, L. Alternative approaches for measuring duration of work disability after low back injury based on administrative workers’ compensation data. Am. J. Ind. Med. 35, 604–618 (1999).

20. Australian Safety and Compensation Council. Type of Occurrence Classification System: Third Edition (Revision 1). (Australian Safety and Compensation Council, 2008).

21. Lopez Bernal, J, Cummins, S & Gasparrini, A. The use of controls in interrupted time series studies of public health interventions. Int. J. Epidemiol. 47, 2082–2093 (2018).

22. Jebb, AT, Tay, L, Wang, W & Huang, Q. Time series analysis for psychological research: examining and forecasting change. Front. Psychol. 6, (2015).

23. Beard, E, Marsden, J, Brown, J, et al. Understanding and using time series analyses in addiction research. Addiction 114, 1866–1884 (2019).

24. Gelman, A, Hill, J & Vehtari, A. Regression and Other Stories. (Cambridge University Press, 2020).

25. Fox, J & Weisberg, S. Time-Series Regression and Generalized Least Squares: An Appendix to An R Companion to Applied Regression, third edition. in An R Companion to Applied Regression 1–8 (2018).

26. Shadish, WR, Cook, T. & Campbell, DT. Experimental and Quasi-Experimental Designs for Generalized Causal Inference. (Houghton Mifflin, 2002).

27. R Core Team. R: A language and environment for statistical computing. (2022).

28. RStudio Team. RStudio: Integrated Development for R. (2022).

29. Bolker, B & Robinson, D. broom.mixed: Tidying Methods for Mixed Models. (2020).

30. Hyndman, RJ, Athanasopolous, G, Bergmeir, C, et al. forecast: Forecasting functions for time series and linear models. (2020).

31. Kassambara, A. ggpubr: ‘ggplot2’ Based Publication Ready Plots. (2020).

32. Firke, S. janitor: Simple Tools for Examining and Cleaning Dirty Data. (2021).

33. Grolemund, G & Wickham, H. Dates and Times Made Easy with lubridate. J. Stat. Softw. 40, 1–25 (2011).

34. Tierney, N, Cook, D, McBain, M & Fay, C. naniar: Data Structures, Summaries, and Visualisations for Missing Data. (2020).

35. Pinheiro, J, Bates, D, DebRoy, S, Sarkar, D, & R Core Team. nlme: Linear and Nonlinear Mixed Effects Models. (2020).

36. Wickham, H & Bryan, J. readxl: Read Excel Files. (2019).

37. Wickham, H, Averick, M, Bryan, J, et al. Welcome to the Tidyverse. J. Open Source Softw. 4, 1686 (2019).

38. Zeileis, A & Grothendieck, G. zoo: S3 Infrastructure for Regular and Irregular Time Series. arXiv:math/0505527 (2005).

39. Lane, T. Effects of South Australia’s 2015 RTW Act on injured worker outcomes. Bridges (2022). Available at: https://doi.org/10.26180/21625778.v1.

40. Lane, TJ. Large-scale workers’ compensation system reform and injured worker outcomes: an interrupted time series. Open Science Framework (2020). Available at: https://osf.io/bwysr/. (Accessed: 11th January 2023)

41. Lane, T. Overhauling a troubled workers’ compensation system: the effects of reform on claim processes and injured worker outcomes. (2020). doi:10.17605/OSF.IO/BWYSR

42. The Social Research Centre. Return to Work Survey 2016 Summary Research Report (Australia & New Zealand). (The Social Research Centre, 2016).

43. Dasinger, LK, Krause, N, Deegan, LJ, Brand, R. & Rudolph, L. Duration of work disability after low back injury: A comparison of administrative and self-reported outcomes. Am. J. Ind. Med. 35, 619–631 (1999).

44. Collie, A, van Vreden, C, Lane, TJ, Sheehan, L. & Iles, RA. Barriers and Enablers for Return to Work: Findings from a Survey of RTW Professionals. (Monash University, 2020).

